# Estimating the infection and case fatality ratio for COVID-19 using age-adjusted data from the outbreak on the Diamond Princess cruise ship

**DOI:** 10.1101/2020.03.05.20031773

**Authors:** Timothy W Russell, Joel Hellewell, Christopher I Jarvis, Kevin Van Zandvoort, Sam Abbott, Ruwan Ratnayake, CMMID COVID-19 working group, Stefan Flasche, Rosalind M Eggo, W John Edmunds, Adam J Kucharski

**Author notes:** authors contributed equally (Joel Hellewell, Christopher I Jarvis, Kevin Van Zandvoort). The members of the Centre for the Mathematical Modelling of Infectious Diseases (CMMID) COVID-19 working group are listed at the end of the article.

## Abstract

Adjusting for delay from confirmation-to-death, we estimated case and infection fatality ratios (CFR, IFR) for COVID-19 on the Diamond Princess ship as 2.3% (0.75%–5.3%) and 1.2% (0.38–2.7%). Comparing deaths onboard with expected deaths based on naive CFR estimates using China data, we estimate IFR and CFR in China to be 0.5% (95% CI: 0.2–1.2%) and 1.1% (95% CI: 0.3–2.4%) respectively.

**Aim:** To estimate the infection and case fatality ratio of COVID-19, using data from passengers of the Diamond Princess cruise ship while correcting for delays between confirmation-and-death, and age-structure of the population.

---

In real-time, estimates of the case fatality ratio (CFR) and infection fatality ratio (IFR) can be biased upwards by under-reporting of cases and downwards by failure to account for the delay from confirmation-to-death. Collecting detailed epidemiological information from a closed population such as the quarantined Diamond Princess can produce a more comprehensive description of asymptomatic and symptomatic cases and their subsequent outcomes. Using data from the Diamond Princess, and adjusting for delay from confirmation-to-outcome and age-structure of the ship’s occupants, we estimated the IFR and CFR for the outbreak in China.

As of 3rd March 2020, there have been 92,809 confirmed cases of coronavirus disease 2019 (COVID-19), with 3,164 deaths [1]. On 1st February 2020, a patient tested positive for COVID-19 in Hong Kong; they disembarked from the Diamond Princess cruise ship on the 25th January [2,3]. This patient had onset of symptoms on the 19th January, one day before boarding the ship [2]. Upon returning to Yokohama, Japan, on February 3rd, the ship was held in quarantine, during which testing was performed in order to measure COVID-19 infections among the 3,711 passengers and crew members onboard.

Passengers were initially to be held in quarantine for 14 days. However, those that had intense exposure to the confirmed case-patient, such as sharing a cabin, were held in quarantine beyond the initial 14-day window [3]. By 20^th^ February, there were 634 confirmed cases onboard (17%), with 328 of these asymptomatic (asymptomatic cases were either self-assessed or tested positive before symptom onset) [3]. Overall 3,063 PCR tests were performed among passengers and crew members. Testing started among the elderly passengers, descending by age [3]. For details on the testing procedure, see [2] and [3].

## Adjusting for outcome delay in CFR estimates

During an outbreak, the so-called naive CFR (nCFR), i.e. the ratio of reported deaths date to reported cases to date, will underestimate the true CFR because the outcome (recovery or death) is not known for all cases [4,5]. We can therefore estimate the true denominator for the CFR (i.e. the number of cases with known outcomes) by accounting for the delay from confirmation-to-death [5].

We assumed the delay from confirmation-to-death followed the same distribution as estimated hospitalisation-to-death, based on data from the COVID-19 outbreak in Wuhan, China, between the 17^th^ December 2019 and the 22^th^ January 2020, accounting right-censoring in the data as a result of as-yet-unknown disease outcomes (Figure 1, panels A and B) [6]. As a sensitivity analysis, we also consider raw “non-truncated” distributions, which do not account for censoring; the raw and truncated distributions have a mean of 8.6 days and 13 days respectively. To correct the CFR, we use the case and death incidence data to estimate the number of cases with known outcomes [5]:

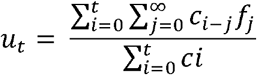

where c_t_ is the daily case incidence at time t, f_t_ is the proportion of cases with delay between onset or hospitalisation and death. u_t_ represents the underestimation of the known outcomes [5] and is used to scale the value of the cumulative number of cases in the denominator in the calculation of the cCFR. Finally, we used the measured proportions of asymptomatic to symptomatic cases on the Diamond Princess to scale the corrected CFR (cCFR) to estimate the infection fatality ratio (IFR).

**Figure 1:**
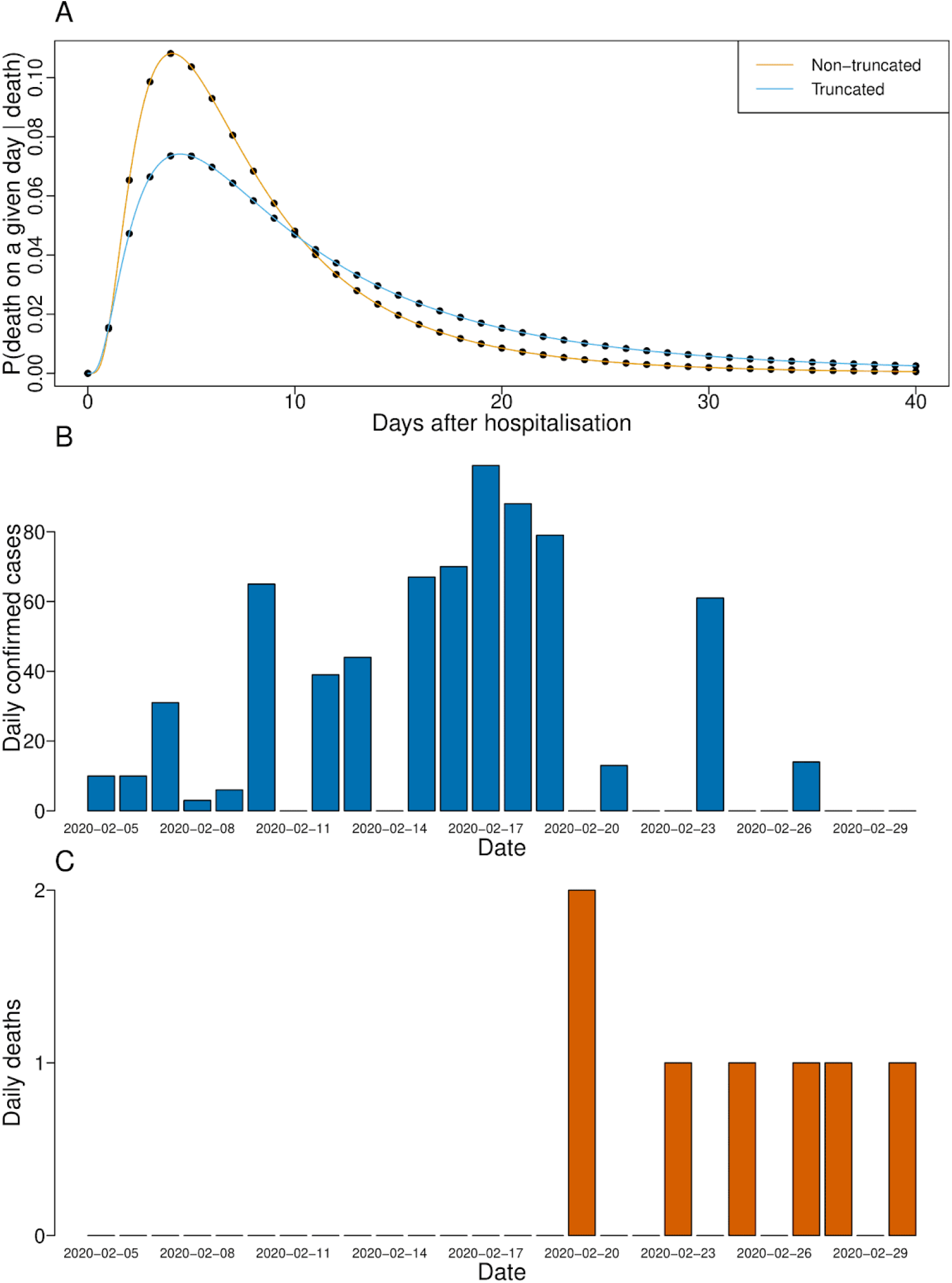
The time-to-death distributions and case and death data used to calculate the cCFR estimates. Panel A: the delay distributions of hospitalisation-to-death; both are lognormal distributions fitted and reported in Linton et al. using data from the outbreak in Wuhan, China. The non-truncated distribution has a mean of 8.6 days and SD of 6.7 days; the right-truncated distribution has a mean of 13 days and SD of 12.7 days. Panels B and C: the case and death timeseries (respectively) of passengers onboard the ship.

## Corrected IFR and CFR estimates

We estimated that the all-age cIFR on the Diamond Princess was 1.2% (0.38–2.7%) and the cCFR was 2.3% (0.75–5.3%) (Table 1). Using the age distribution of cases and deaths on the ship [2,3], we estimated that for individuals aged 70 and over, the cIFR was 9.0% (3.8–17%) and the cCFR was 18% (7.3–33%) (Table 1). The 95% confidence intervals were calculated with an exact binomial test with death count and either cases or known outcomes (depending on whether it was an interval for the naive or corrected estimate).

**Table 1:**
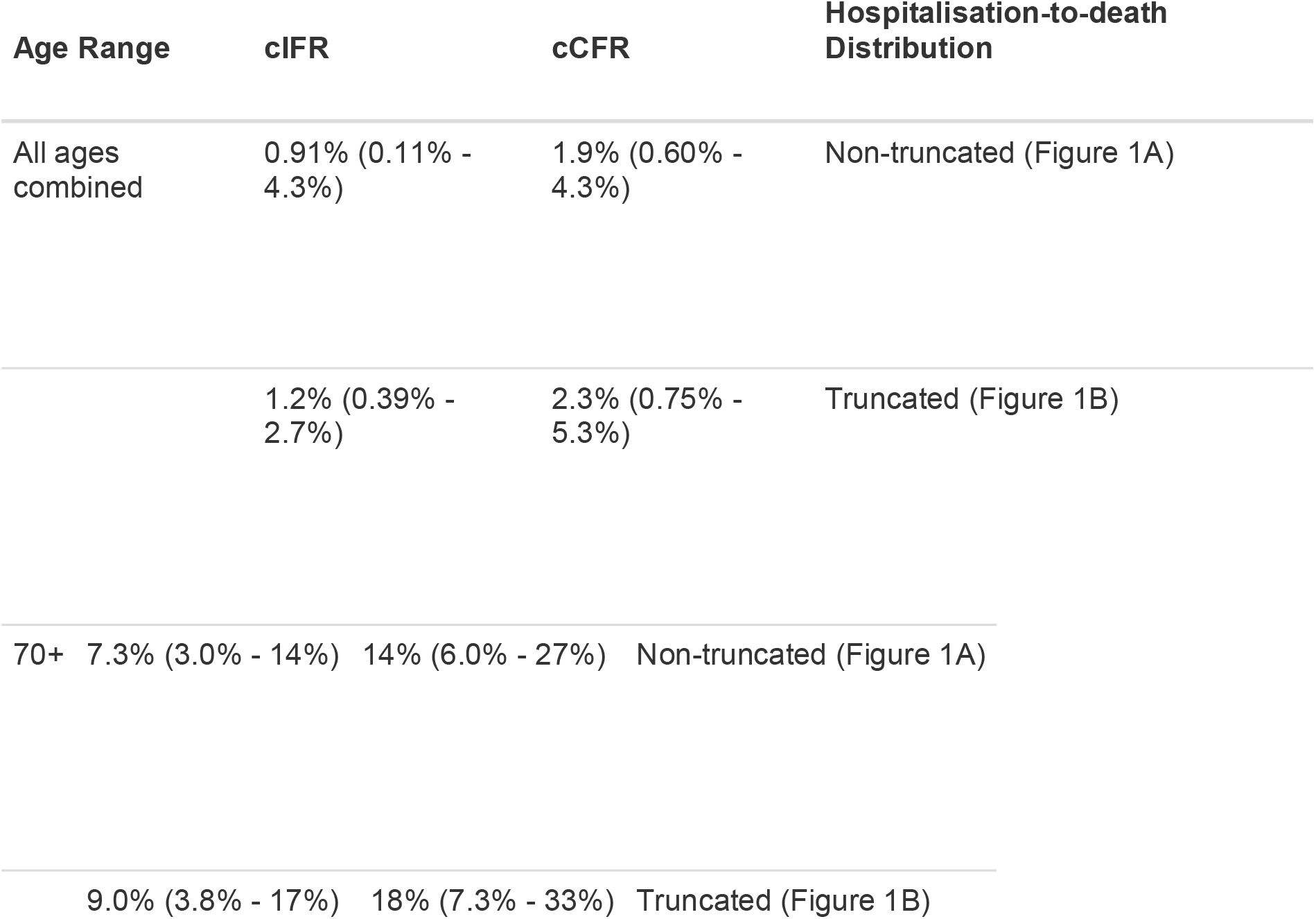
cIFR and cCFR estimates calculated using the reported case and death data on the Diamond Princess cruise ship [2]. Correction was performed using equation (1) and the hospitalisation-to-death distribution in [6].

| Age Range | cIFR | cCFR | Hospitalisation-to-death Distribution |
| --- | --- | --- | --- |
| All ages combined | 0.91% (0.11% - 4.3%) | 1.9% (0.60% - 4.3%) | Non-truncated (Figure 1A) |
|  | 1.2% (0.39% - 2.7%) | 2.3% (0.75% - 5.3%) | Truncated (Figure 1B) |
| 70+ | 7.3% (3.0% - 14%) | 14% (6.0% - 27%) | Non-truncated (Figure 1A) |
|  | 9.0% (3.8% - 17%) | 18% (7.3% - 33%) | Truncated (Figure 1B) |

Using the age-stratified nCFR estimates reported in a large study in China [7], we then calculated the expected number of deaths of people who were onboard the ship in each age group, assuming this nCFR estimate was accurate. This produced a total of 15.15 expected deaths, which gives a nCFR estimate of 5% (15.15/301) for Diamond Princess (Table 2), which falls within the top end of our 95% CI. As our corrected cCFR for Diamond Princess was 2.3% (0.75% - 5.3%), this suggests we need to multiply the nCFR estimates in China [7] by a factor 46% (95% CI: 15–105%) to obtain the correct value. As the raw overall nCFR reported in the China data was 2.3% [7], this suggests the cCFR in China during that period was 1.1% (95% CI: 0.3-2.4%) and the IFR was 0.5% (95% CI: 0.2-1.2%). Based on cases and deaths reported in China up to 4th March 2020, nCFR = 2984/80422*100 = 3.71% (95% CI 3.58% - 3.84%); this naive value is significantly higher than the corrected CFR we estimate here.

**Table 2:** Age stratified data of symptomatic (symp.) and asymptomatic (asymp.) cases on-board the Diamond Princess [2], [3], along with the nCFR estimates given in [7], the expected number of cases in each age group if the nCFR estimates were correct where the total number of expected deaths under these estimates was 15.15 and age stratified observed/expected death ratios.

| Age Range | No. of passengers | Symp. cases | Asymp. cases | nCFR | Expected deaths using nCFR | Observed deaths on cruise ship |
| --- | --- | --- | --- | --- | --- | --- |
| 0 - 9 | 16 | 0 | 1 | 0.0%<br>(0.0% - 0.9%) | 0 (0 - 0) | 0 |
| 10 - 19 | 23 | 2 | 3 | 0.2%<br>(0.0% - 1.0%) | 0 (0 - 0) | 0 |
| 20 - 29 | 347 | 25 | 3 | 0.2%<br>(0.1% - 0.4%) | 0.05 (0.02 - 0.10) | 0 |
| 30 - 39 | 428 | 27 | 7 | 0.2%<br>(0.1% - 0.4%) | 0.06 (0.04 - 0.10) | 0 |
| 40 - 49 | 334 | 19 | 8 | 0.4%<br>(0.3% - 0.6%) | 0.08 (0.06 - 0.12) | 0 |
| 50 - 59 | 398 | 28 | 31 | 1.3%<br>(1.1% - 1.5%) | 0.36 (0.31 - 0.43) | 0 |
| 60 - 69 | 923 | 76 | 101 | 3.6%<br>(3.2% - 4.0%) | 2.74 (2.5 - 3.1) | 0 |
| 70 - 79 | 1015 | 95 | 139 | 8.0%<br>(7.2% - 8.9%) | 7.6 (6.8 - 8.4) | 6 |
| 80 - 89 | 216 | 29 | 25 | 14.8%<br>(13.0% - 16.7%) | 4.28 (3.8 - 4.9) | 1 |
| Totals | 3711 | 301 | 318 |  | 15.15 (13.5 - 17.1) | 7 |

The case fatality ratio is challenging to accurately estimate in real time [8], especially for an infection with attributes similar to SARS-CoV-2, which has a delay of almost two weeks between confirmation and death, strong effects of age-dependence and comorbidities on mortality risk, and likely under-reporting of cases in many settings [6]. Using an age-stratified adjustment, we accounted for changes in known outcomes over time. By applying the method to Diamond Princess data, we focus on a setting that is likely to have lower reporting error because large numbers were tested and the test has high sensitivity.

The average age onboard the ship was 58, so our estimates of cCFR cannot directly be applied to a younger population; we therefore scaled our estimates to obtain values for a population equivalent to those in the early China outbreak. There were some limitations to our analysis. Cruise ship passengers may have a different health status to the general population of their home countries, due to health requirements to embark on a multi-week holiday, or differences related to socio-economic status or comborbities. Deaths only occurred in individuals 70 years or older, so we were not able to generate age-specific cCFRs; the fatality risk may also be influenced by differences in healthcare between countries. Because of likely age-specific differences in reporting, we focused on overall cCFR in China, rather than calculating age-specific cCFRs [7].

## Data Availability

All of the data and the code required to reproduce the figures and results of this study can be found at the public github repository: https://github.com/thimotei/cCFRDiamondPrincess.

https://github.com/thimotei/cCFRDiamondPrincess.

## Author contributions

TWR, AJK and WJE conceived of the study and collected the data. TWR, AJK and SA coded the methods. TWR and JH wrote the first draft of the manuscript with feedback from all other authors. KvZ, TWR, SA and CIJ worked on the statistical aspects of the study. All authors read and approved the final version of the manuscript. Each member of the CMMID COVID-19 working group contributed in processing, cleaning an interpretation of data, interpreted findings, contributed to the manuscript, and approved the work for publication.

## Acknowledgements

TWR, JH SA, SF and AJK are supported by the Wellcome Trust (grant numbers: 206250/Z/17/Z, 210758/Z/18/Z, 210758/Z/18/Z, 210758/Z/18/Z, 208812/Z/17/Z, 206250/Z/17/Z). CIJ is supported by Global Challenges Research Fund (GCRF) project ‘RECAP’ managed through RCUK and ESRC (ES/P010873/1). KvZ is supported by Elrha’s Research for Health in Humanitarian Crises (R2HC) Programme, which aims to improve health outcomes by strengthening the evidence base for public health interventions in humanitarian crises. The R2HC programme is funded by the UK Government (DFID), the Wellcome Trust, and the UK National Institute for Health Research (NIHR). RR is supported by Canadian Institutes of Health Research (Award no. DFS-164266). RME is supported by HDR UK (grant: MR/S003975/1)

CMMID nCoV working group funding statements: Thibaut Jombart (RCUK/ESRC (grant: ES/P010873/1); UK PH RST; NIHR HPRU Modelling Methodology), Amy Gimma (GCRF (ES/P010873/1)), Nikos I Bosse (no funding statement to declare), Alicia Rosello (NIHR (grant: PR-OD-1017-20002)), Mark Jit (Gates (INV-003174), NIHR (16/137/109)), James D Munday(Wellcome Trust (grant: 210758/Z/18/Z)), Billy J Quilty (NIHR (16/137/109)), Petra Klepac (Gates (INV-003174)), Hamish Gibbs (NIHR (ITCRZ 03010)), Yang Liu (Gates (INV-003174), NIHR(16/137/109)), Sebasitan Funk (Wellcome Trust (grant: 210758/Z/18/Z)), Samuel Clifford (Wellcome Trust (grant: 208812/Z/17/Z)), Fiona Sun (NIHR EPIC grant (16/137/109)), Kiesha Prem (Gates (INV-003174)), Charlie Diamond (NIHR (16/137/109)), Nicholas Davies (NIHR (HPRU-2012-10096)), Carl A B Pearson

## Members of the Centre for the Mathematical Modelling of Infectious Diseases (CMMID) nCoV working group

The following authors were part of the Centre for Mathematical Modelling of Infectious Disease 2019-nCoV working group. Thibaut Jombart, Amy Gimma, Nikos I Bosse, Alicia Rosello, Mark Jit, James D Munday, Billy J Quilty, Petra Klepac, Hamish Gibbs, Yang Liu, Sebasitan Funk, Samuel Clifford, Fiona Sun, Kiesha Prem, Charlie Diamond, Nicholas Davies, Carl A B Pearson.

## Conflict of interest

None declared.

## Notes

### Competing Interest Statement

The authors have declared no competing interest.

